# Understanding ‘Second Victim Experience’ among staff at Kijabe Hospital

**DOI:** 10.1101/2025.06.28.25330416

**Authors:** Faith Lelei, Moses Odhiambo Osoo, Mary B. Adam, Elijah Ntomariu, Peris Kiarie

**Author notes:** **Correspondence**: Faith Lelei, P.O Box 20-00220 Kijabe, Kenya. |.

## Abstract

**Introduction:** Patient safety is crucial in healthcare. However, medical errors still occur and can harm patients. When healthcare workers experience adverse events, they can become “second victims” and experience emotional turmoil. Symptoms can persist for months and include hypervigilance and doubts regarding clinical skills. We conducted a study among staff at Kijabe Hospital to examine the second victim experience (SVE) and develop support strategies.

**Methods:** We conducted an online survey using the Second Victim Experience and Support Tool in May 2023.

**Results:** Of 900 healthcare workers,121 (13.4%) participated in the survey. The respondents consisted of various cadres including 31 students, 24 nurses, 18 clinical officers, 13 allied health workers, 12 consultants, 10 residents, and 13 from other cadres. Overall, 67.2% of the participants reported an SVE. Among these, 89.7% occurred within the past 12 months of these study. Of these participants, 49.6% provided hands-on or direct patient care, and 38.0% provided both direct and indirect patient care. The psychosocial symptoms reported included depressed mood (27.3%), guilt and frustration (27.3%), feelings of inadequacy and loneliness (24.0%), irritability (20.7%), and recurrent intrusive thoughts or images (19.8%). The physical symptoms reported included fatigue (24.0%), headaches (14.9%), and sleep disturbances (11.6%). Forty-five percent of SVs received support from someone at the hospital, while 27.3% received support from a colleague or a peer. In addition, 22.3% received support from close friends or family members, 6.7% received support from hospital administration or management and 0.8% received support from a pastor and an external counselor.

**Conclusions:** Two-thirds of healthcare workers had a second victim experience, reporting fatigue, depressed mood, guilt, and frustration. Counseling and psychological support were preferred by staff. Hospital management needs to prioritize staff support and acknowledge this experience to ensure patient safety.

## Introduction

Patient safety is becoming increasingly important as healthcare systems continue to be more complex. This requires more of healthcare workers in terms of both knowledge and skills. While patient safety is a paramount issue for all healthcare workers, unfortunately, medical errors occur resulting in events that put patients at harm. Healthcare workers often respond emotionally to these events. The term second victim was first used by A. W. Wu in a 2000 British Journal of Medicine article where he described the experience that has been termed “second victim”; “Virtually every practitioner knows the sickening realization of making a bad mistake. You feel singled out and exposed—seized by the instinct to see if anyone has noticed it. You agonize about what to do, whether to tell anyone, and what to say. Later, the event replays itself repeatedly in your mind(1), and Dr. Wu goes on to say that a healing mechanism is needed for the healthcare worker.

Extensive evidence has been obtained since landmark papers on the emotional turmoil and longer-term influence of patient safety incidents on the health care worker, “the second victim’(2–6). A consensus definition of second victim reported from Scott et al is, “A second victim is a health care provider involved in an unanticipated adverse patient event, medical error and/or a patient-related injury who become victimized in the sense that the provider is traumatized by the event. Frequently, second victims feel personally responsible for the unexpected patient outcomes and feel as though they have failed their patients, second-guessing their clinical skills and knowledge base.”(7).

The literature describes both the psychological and psychosomatic symptoms of healthcare workers who are second victims(8,9). Symptoms persisted for 1-3 months in many providers and ranged from hypervigilance (53%) to doubts about the knowledge and skills of the team (27%). Symptoms were 8-fold more likely to persist when the patient died as a result of the incident. Second, victims develop a variety of coping mechanisms(10) and report desiring support. Literature describing treatment and support services, as well as tools organizations can utilize, has been published (2,11–13), demonstrating that there is a recovery process for those who suffer from the symptoms described above (7,14,15).

Patient safety is compromised when there are medical errors and this has also been studied in developing countries. Wilson et al. examined records from 8 countries, including Kenya, and sampled 26 hospitals. Records from 15,548 patients were reviewed, and more than 8% of patients had an error identified. It was estimated that 83% of these errors were preventable and 30% of the errors resulted in patient death (16). This means that a sizable number of healthcare workers in Africa are at risk of experiencing the symptoms of a second victim. These health care workers may represent what Scott et al call “the walking wounded” yet there is scanty literature about extent of second victim experience in health care professionals in sub–Saharan Africa. The purpose of this study was to examine the second victim experience of the staff at Kijabe Hospital. The long-term goal of this study was to develop support strategies for the staff at Kijabe Hospital.

## Materials and Methods

### Study Design

This was a cross-sectional online survey conducted using the validated Second Victim Experience and Support Too (11) deployed in an online anonymous survey in Redcap. The study was open to all employed staff at Kijabe Hospital and trainees who were part of the clinical division, regardless of their cadre or level of training. The survey was conducted anonymously, and staff members were invited to participate through official hospital channels including chapels, department-specific awareness sessions, WhatsApp groups, and email channels in May 2023.

### Setting

AIC Kijabe Hospital has served the poor and vulnerable in Kenya for more than 100 years. It is a 330-bed facility that performs more than 8,500 surgical procedures per year and sees over 130,000 outpatients with a preponderance of women and children as its patient population. The institution has grown from a small mission outpost to a major tertiary care hospital and training center. It is an internship center for clinical, nutritional, and medical officers with multiple residency programs. Kijabe College of Health Sciences has graduated from nurses for over 40 years.

The institution currently trains healthcare professionals at various levels, including nurses, clinical officers, perioperative theatre technicians, several higher diploma and residency programs.

### Participants

All Kijabe Hospital staff and trainees at any level were invited to participate in the survey.

### Briefing of Participants

Participants were provided detailed briefings before and during recruitment through department meetings, chapel sessions, and digital communication (email and WhatsApp), clearly outlining the study’s purpose, confidentiality measures, voluntary nature, and emotional support options. Completion of the anonymized REDCap survey was considered evidence of informed consent at the individual level. This process, and the absence of personal identifiers, was designed to protect autonomy and reduce potential coercion or social desirability bias, with opportunities for clarification and support provided in real time and via contact information for the hospital psychology team.

### Sample size

Response rates from healthcare workers have consistently been low, typically ranging between 10-15%. Although we aimed for a 20% sample size of the total staff, only 171 staff members attempted the survey, and out of those, 121 provided answers regarding their experiences as second victims. This resulted in a response rate of 19% (171 out of 900).

### Data Collection

All data was collected electronically through a redcap survey link. The data was only accessible to study personnel. All the variables collected were categorical

### Data Analysis

The demographic characteristics of the participants, their experiences as second victims, reported physical and psychosocial symptoms, supportive strategies, and support mechanisms were assessed using descriptive statistics. Specifically, the frequency, percentages, and rank order of responses were used to analyze the data.

only 171 staff members attempted the survey, and out of those, 121 provided answers regarding their experiences as second victims.

Missing data for 50 healthcare workers without SVE data were dropped from subsequent analysis through pairwise deletion

The analysis was conducted using STATA version 18.

### Ethical Considerations

The study was approved by the Institutional Scientific and Ethical Review Committee (ISERC Approval No: KH/ISERC/02718/0036/2022)

### Data Availability

Availability of data and materials will be provided by the corresponding author upon request.

### Funding

This study did not receive any external funding.

### Data confidentiality

Confidentiality was embedded within the online platform, as staff were not identified in any way. The Redcap Survey link did not collect email addresses, phone numbers, or any other identifiable information. The open screen also reminded people where additional emotional or psychological support could be obtained.

## Results

### Participant’s demographic characteristics

Of the 900-hospital staff at the time of this study, 121 (13.4%) participated in the survey. The sample consisted of multiple cadres of healthcare workers and trainees. Students (including Diploma and Higher diploma levels) were 25.6%, licensed nurses 19.8%, clinical officers 14.9%, allied health 10.7%, consultants 9.9%, residents and medical officers 8.3%, and one patient assistant. Of these participants, 38.8% had worked in healthcare for 1–5 years, 28.1% for 6–10 years, and 6.6% for more than 16 years. The majority of participants (49.5%) were involved in hands-on or direct patient care, whereas 38.7% were involved in both direct and indirect patient care. (Table 1)

**Table 1.** Participants demographics characteristics’.

| Participants Characteristics' | Total (N=121, %) |
| --- | --- |
| Please check off your main role at KH, n (%) |  |
| Allied Health: Physiotherapy, OT, Audiology, Pharmacy, Nutrition, Lab, Radiology | 13 (10.7) |
| Consultants | 12 (9.9) |
| Residents & MOs | 10 (8.3) |
| Registered Clinical Officer and specialized Clinical Officers (ECCCO, PECCCO) | 18 (14.9) |
| Nurse & Nursing Specialties (e.g. Critical Care, Renal, KRNA) | 24 (19.8) |
| Patient Assistants & surgical technologists | 1 (0.8) |
| Students (Nursing, Clinical officers, ECCCO, PECCCO, FHCO, KRNA, KRCCN, POTT) | 31 (25.6) |
| Interns (Medical Officer, Clinical officer, Nutritionist) | 4 (3.3) |
| Admin Team, Human Resource, Finance Team & Information communication technology | 5 (4.2) |
| Support Team (Catering, security/Keeping, Laundry) | 3 (2.5) |
| How long have you worked in health care? n (%) |  |
| Less than 1 year | 19 (15.7) |
| 1 - 5 years | 47 (38.8) |
| 6 - 10 years | 34 (28.1) |
| 11 - 15 years | 13 (10.8) |
| More than 16 years | 8 (6.6) |
| At the hospital I provide, n (%) |  |
| Hands-on or direct patient care | 59 (49.5) |
| Indirect patient care or in support of patient care (managing, educating, developing, implementing, delivering, administering the business of the hospital, e.g., finance, materials management etc.) | 8 (6.7) |
| Provide both direct patient care and indirect patient care | 46 (38.7) |
| Provide neither direct or indirect patient care | 2 (1.7) |
| Other | 4 (3.4) |

*ECCCO- Emergency and Critical Care Clinical Officer, PECCCO- Paediatric Emergency and Critical Care, FHCO-Family Health Clinical officer, OT- Occupational Therapy, KRNA-Kenya Registered Nurse Anaesthetist, KRCCN -Kenya Registered Critical Care Nursing, POTT-Perioperative theatre technician*

### Second victim experience among healthcare workers at Kijabe hospital

Overall, 67.2% of the participants reported to have had a second victim experience (SVE). Among these, 89.7% occurred within the past 12 months. Notably 48.7% of the SVE cases provided hands-on or direct patient care, whereas 39.7% provided both direct and indirect patient care. SVE was reported highest among students (25.6%), followed by nurses and nursing specialties (24.4%), Clinical Officers and specialized clinical officers (16.7%), consultants (14.1%), and Residents & Medical officers (10.3%). Many of the participants who experienced SVE had worked in healthcare for 1–5 years (37.9%), followed by those who had worked for 6–10 years (28.4%). Participants who had worked for more than six years had higher cases of SVE than those who had worked for fewer years. For instance, of the eight participants who had worked in healthcare for more than 16 years, seven (87.5%) had SVE, the highest compared to those who had worked for less than 16 years. In terms of departments, SVE was reported by 33.3% of participants working in the Adult Medical-Surgical ward, BKKH (Pediatric ward, NICU, PICU, and Nursery), 30.8% and 24.4% in Theatre, 24.4% in MCH and OPD, 21.8% in ICU, 19.2% in maternity, and 19.2% in other departments, as shown in Table 2.

**Table 2:**
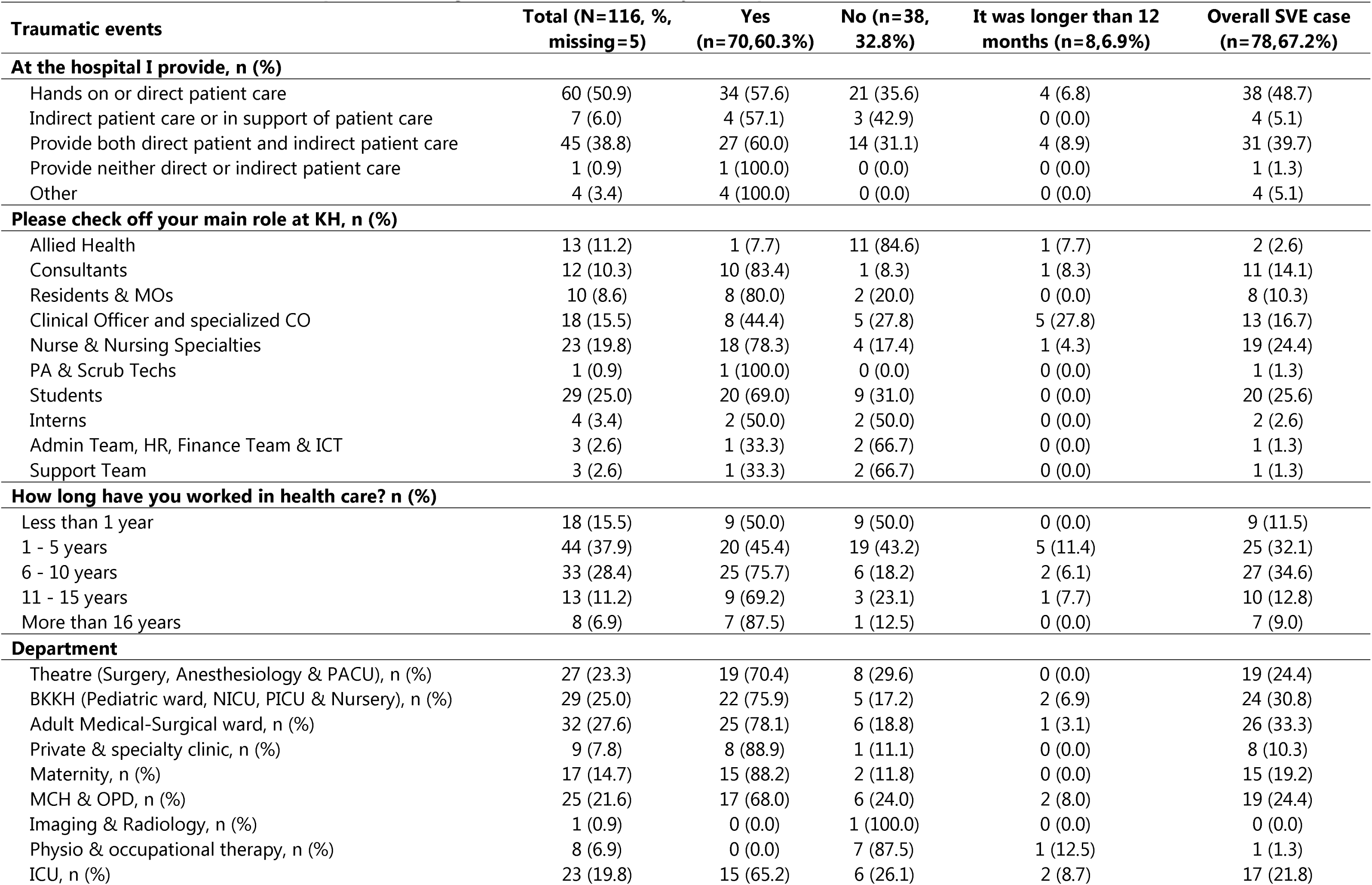
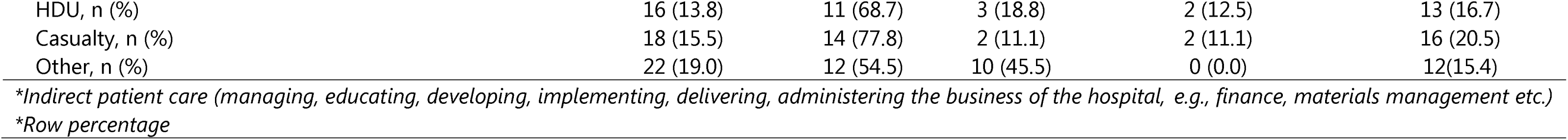
Second victim experience among healthcare workers at Kijabe hospital atic events.

| **Traumatic events** | **Total (N=116, %, missing= 5)** | **Yes (n=70,60.3%)** | **No (n=38, 32.8%)** | **It was longer than 12 months (n=8,6.9%)** | **Overall SVE case (n=78,67.2%)** |
| --- | --- | --- | --- | --- | --- |
| **At the hospital I provide, n (%)** |  |  |  |  |  |
| Hands on or direct patient care | 60 (50.9) | 34 (57.6) | 21 (35.6) | 4 (6.8) | 38 (48.7) |
| Indirect patient care or in support of patient care | 7 (6.0) | 4 (57.1) | 3 (42.9) | 0 (0.0) | 4 (5.1) |
| Provide both direct patient and indirect patient care | 45 (38.8) | 27 (60.0) | 14 (31.1) | 4 (8.9) | 31 (39.7) |
| Provide neither direct or indirect patient care | 1 (0.9) | 1 (100.0) | 0 (0.0) | 0 (0.0) | 1 (1.3) |
| Other | 4 (3.4) | 4 (100.0) | 0 (0.0) | 0 (0.0) | 4 (5.1) |
| **Please check off your main role at KH, n (%)** |  |  |  |  |  |
| Allied Health | 13 (11.2) | 1 (7.7) | 11 (84.6) | 1 (7.7) | 2 (2.6) |
| Consultants | 12 (10.3) | 10 (83.4) | 1 (8.3) | 1 (8.3) | 11 (14.1) |
| Residents & MOs | 10 (8.6) | 8 (80.0) | 2 (20.0) | 0 (0.0) | 8 (10.3) |
| Clinical Officer and specialized CO | 18 (15.5) | 8 (44.4) | 5 (27.8) | 5 (27.8) | 13 (16.7) |
| Nurse & Nursing Specialties | 23 (19.8) | 18 (78.3) | 4 (17.4) | 1 (4.3) | 19 (24.4) |
| PA & Scrub Techs | 1 (0.9) | 1 (100.0) | 0 (0.0) | 0 (0.0) | 1 (1.3) |
| Students | 29 (25.0) | 20 (69.0) | 9 (31.0) | 0 (0.0) | 20 (25.6) |
| Interns | 4 (3.4) | 2 (50.0) | 2 (50.0) | 0 (0.0) | 2 (2.6) |
| Admin Team, HR, Finance Team & ICT | 3 (2.6) | 1 (33.3) | 2 (66.7) | 0 (0.0) | 1 (1.3) |
| Support Team | 3 (2.6) | 1 (33.3) | 2 (66.7) | 0 (0.0) | 1 (1.3) |
| **How long have you worked in health care? n (%)** |  |  |  |  |  |
| Less than 1 year | 18 (15.5) | 9 (50.0) | 9 (50.0) | 0 (0.0) | 9 (11.5) |
| 1 - 5 years | 44 (37.9) | 20 (45.4) | 19 (43.2) | 5 (11.4) | 25 (32.1) |
| 6 - 10 years | 33 (28.4) | 25 (75.7) | 6 (18.2) | 2 (6.1) | 27 (34.6) |
| 11 - 15 years | 13 (11.2) | 9 (69.2) | 3 (23.1) | 1 (7.7) | 10 (12.8) |
| More than 16 years | 8 (6.9) | 7 (87.5) | 1 (12.5) | 0 (0.0) | 7 (9.0) |
| **Department** |  |  |  |  |  |
| Theatre (Surgery, Anesthesiology & PACU), n (%) | 27 (23.3) | 19 (70.4) | 8 (29.6) | 0 (0.0) | 19 (24.4) |
| BKKH (Pediatric ward, NICU, PICU & Nursery), n (%) | 29 (25.0) | 22 (75.9) | 5 (17.2) | 2 (6.9) | 24 (30.8) |
| Adult Medical-Surgical ward, n (%) | 32 (27.6) | 25 (78.1) | 6 (18.8) | 1 (3.1) | 26 (33.3) |
| Private & specialty clinic, n (%) | 9 (7.8) | 8 (88.9) | 1 (11.1) | 0 (0.0) | 8 (10.3) |
| Maternity, n (%) | 17 (14.7) | 15 (88.2) | 2 (11.8) | 0 (0.0) | 15 (19.2) |
| MCH & OPD, n (%) | 25 (21.6) | 17 (68.0) | 6 (24.0) | 2 (8.0) | 19 (24.4) |
| Imaging & Radiology, n (%) | 1 (0.9) | 0 (0.0) | 1 (100.0) | 0 (0.0) | 0 (0.0) |
| Physio & occupational therapy, n (%) | 8 (6.9) | 0 (0.0) | 7 (87.5) | 1 (12.5) | 1 (1.3) |
| ICU, n (%) | 23 (19.8) | 15 (65.2) | 6 (26.1) | 2 (8.7) | 17 (21.8) |
| HDU, n (%) | 16 (13.8) | 11 (68.7) | 3 (18.8) | 2 (12.5) | 13 (16.7) |
| Casualty, n (%) | 18 (15.5) | 14 (77.8) | 2 (11.1) | 2 (11.1) | 16 (20.5) |
| Other, n (%) | 22 (19.0) | 12 (54.5) | 10 (45.5) | 0 (0.0) | 12(15.4) |

*\*Indirect patient care (managing, educating, developing, implementing, delivering, administering the business of the hospital, e.g., finance, materials management etc.)*
*\*Row percentage*

### Reported physical and psychosocial symptoms after SVE among Healthcare workers at Kijabe Hospital

Fatigue was the most common physical symptom reported by 24% of the participants, followed by headaches (14.9%) and trouble sleeping or oversleeping (11.6%). Additionally, four participants indicated drug and alcohol abuse. The most commonly reported psychosocial symptoms were depressed mood (27.3%), guilt and frustration (27.3%) and feelings of inadequacy and loneliness (24.0%). Other symptoms reported were irritability (20.7%), recurrent images or thoughts of the event (19.8%), a strong need to talk about the event or read (19.8%), loss of interest or pleasure (16.5%), hypervigilance with everything one does (16.5%), and feeling distracted (16.5%). Triggers due to non-specific events and the desire to connect with others experiencing similar trauma was each reported by 7 participants. (Table 3)

**Table 3.** Reported physical and psychosocial symptoms after SVE among Healthcare workers at Kijabe Hospital. Most commonly reported physical and psychosocial symptoms.

| **Physical symptoms** | **n, %** | **Psychosocial symptoms** | **n, %** |
| --- | --- | --- | --- |
| Fatigue | 29(24.0) | Depressed mood | 33(27.3) |
| Headaches | 18(14.9) | Guilt & frustration | 33(27.3) |
| Trouble Sleeping or Sleeping too much | 14(11.6) | Feelings of inadequacy and loneliness | 29(24.0) |
| Trouble eating | 9(7.4) | Irritability | 25(20.7) |
| Less activity | 5(4.1) | Recurrent images or thoughts of the event | 24(19.8) |
| Drug or alcohol abuse | 4(3.3) | Strong need to talk about the event or read | 24(19.8) |
|  |  | Loss of interest or pleasure | 20(16.5) |
|  |  | Hyper-vigilance with everything you do | 20(16.5) |
|  |  | Feeling distracted | 20(16.5) |
|  |  | Inability to think or concentrate | 19(15.7) |
|  |  | Social isolation | 16(13.2) |
|  |  | Distress when you are exposed to events that Remind you of the trauma | 13(10.7) |
|  |  | Worry about losing job | 11(9.1) |
|  |  | Loss of trust | 10(8.3) |
|  |  | Perceived indifference from colleagues | 10(8.3) |
|  |  | Triggered by non-specific events | 7(5.8) |
|  |  | Desire to connect with others experiencing similar trauma | 7(5.8) |

\* Participants were allowed to select one or more responses.

### Supportive strategies after SVE among Healthcare workers at Kijabe Hospital

Support strategies suggested by most of the participants were access to counseling, psychological, or psychiatric services, whether on-site or off-site (61.2%), followed by the opportunity to take time out of one’s immediate clinical duties to regroup (47.1%). Supportive guidance and mentoring during clinical duties were identified as important by 42.1% of respondents. A safe space to contribute insights on preventing similar events in the future was seen as important by 41.3% of the respondents. Debriefing, crisis intervention, and stress management, either individually or in groups/teams, were identified as important by 40.5% of the respondents. The ability to discuss ethical concerns related to the event or the processes followed subsequently was identified as important by 37.2% of the respondents. Clear and timely information about the processes followed after serious adverse events, such as peer review committees, root cause analyses, and incident reports, was reported as important by 36.4% of the respondents. Formal emotional support from peers was identified as important by 33.1% of respondents. Assistance with communicating with patients and/or families was identified as important by 29.8% of the respondents, as was informal emotional support.

Guidance on the roles expected in the processes following serious adverse events was identified as important by 24.8% of respondents, and help in preparing to participate in these processes was identified as important by 16.5% of respondents, as shown in Table 4.

**Table 4.** Supportive strategies after SVE among Healthcare workers at Kijabe Hospital.

| Supportive strategies | (N=121, %) |
| --- | --- |
| Access to counseling, psychological or psychiatric services (in the work area or off site) | 74(61.2) |
| An opportunity to take time out from your immediate clinical duties to regroup | 57(47.1) |
| Supportive guidance/mentoring as you continued with your clinical duties | 51(42.1) |
| A safe opportunity to contribute any insights you had into how similar events could be prevented in the future. | 50(41.3) |
| Prompt debriefing, crisis intervention stress management (either for individual or for group/team) | 49(40.5) |
| An opportunity to discuss any ethical concerns you had relating to the event or the processes that were followed subsequently. | 45(37.2) |
| Clear and timely information about the processes that are followed after serious adverse events (e.g. peer review committees, root cause analyses, preparation of incident reports) | 44(36.4) |
| Formal emotional support (peer to peer support) | 40(33.1) |
| Help to communicate with the patient and/or family | 36(29.8) |
| Informal emotional support | 36(29.8) |
| Guidance about the roles you were expected to play in the processes that are followed after serious adverse events. | 30(24.8) |
| Help to prepare to participate in the processes that were followed after the serious adverse event. | 20(16.5) |

\* Participants were allowed to select one or more response.

### Support mechanism or help used in processing an adverse event among healthcare workers at Kijabe Hospital

Many participants worked through these traumatic events emotionally on their own (44.6%), and other support mechanisms utilized during these experiences included seeking help outside the hospital (14.9%). Sadly, 9.9% of these participants contemplated leaving the profession of the hospital as a support mechanism to forget their traumatic effects that occurred to them. A relatively low number of patients sought help within their work units (4.3%) or within the hospital (1.9%). Concerning who supported these second victims the most, it is noteworthy that they trusted their colleagues and peers (20.5%), followed by close friends (16.8%), and family members (13.0%). Only eight participants sought help from the administration and/or manager. One participant sought help from a pastor, while the other sought help from an external counselor. One participant did not share this with anyone., respectively, as indicated in table 5.

**Table 5.** Support mechanism or help used in processing an adverse event with 121 who took survey.

| Support mechanism or help used in processing an adverse event | (N=121, %) |
| --- | --- |
| I worked it out emotionally on my own. | 54(33.5) |
| I sought help from outside the hospital. | 18(11.2) |
| I seriously contemplated leaving the profession or the hospital | 12(7.5) |
| I sought help offered within my work unit | 7(4.3) |
| I sought help from the hospital. | 3(1.9) |
| Who supported you following this event |  |
| Colleague/Peer | 33(20.5) |
| Close friend | 27(16.8) |
| Family Member | 21(13.0) |
| Significant other | 10(6.2) |
| Administration/Manager | 8(4.9) |
| I did not share | 1(0.6) |
| Pastor | 1(0.6) |
| External counselling | 1(0.6) |

\* Participants were allowed to select one or more response..

## Discussion

About two-thirds of this study participants had a second victim experience. Our study found an SVE prevalence among staff and trainees with 67.2%, experiencing symptoms within the previous 12 months. The study was conducted post-COVID, a period which was particularly stressful for healthcare workers worldwide and may have influenced staff toward greater reporting. The 67% prevalence was within the range reported in other studies. In a systematic review by Nydoo et al.(17), the authors reported a prevalence of second victim experiences between 10% and 76% within the specialty of obstetrics. However, a study conducted among German physicians by Strametz et al.(18) reported a second victim experience prevalence of 59%, with 35% of them experiencing symptoms in the last 12 months, a lower prevalence than what we found. A study by Rivera-Chiauzzi et al. (19) sought to understand Second Victim Experience among Multidisciplinary Providers in OBGYN, similar to our approach, in the sense that it included both clinical and non-clinical staff members and reported a prevalence of 44.8%, with 18.8% of them experiencing symptoms in the last 12 months, which was lower than our study but this study was also conducted pre-COVID period.

Nydoo et al. and Mathebula et al. (17,20) identified the most common physical and psychological symptoms experienced by healthcare workers who had experienced a traumatic event. As in our study, the most prevalent physical symptoms included fatigue and headaches, while the psychological symptoms included depressed mood, feelings of guilt, loneliness, and feelings of inadequacy. It is important to note that these symptoms have consistently been reported in different settings and studies. In a study conducted by Mathebula et al.(20), more staff members reported experiencing psychological symptoms than physical symptoms, which differs from the findings of this study. This further highlights the importance of not only increasing awareness of the second victim experience, but also providing accessible psychological support to healthcare workers who have experienced traumatic events. Our study found that staff who reported SVE primarily processed the event by themselves, with only a few seeking supports from their managers or supervisors. This is similar to the findings of the study conducted by Strametz et al.(18) however, in the study conducted by Mathebula et al.(20), participants expressed a desire to discuss the event with their managers or supervisors. It is important to explore the reasons why very few healthcare workers seek support from their managers or supervisors.

The impact of SVE on healthcare providers is a complex issue that requires attention from management and hospital leadership. Future studies in these regions could further explore the subject, particularly to understand coping mechanisms and how staff would like hospital leadership and management to support them during such events. Moreover, it may be useful to investigate the potential correlations between staff sick leave and second-victim experiences. This could provide a deeper understanding of how SVE can impact healthcare providers and inform the development of appropriate support systems to help them cope with the aftermath.

It is important to highlight that error reporting is beginning to take shape in Africa, primarily within the context of pharmacovigilance (21–24). Hospitals should strive to create an enabling environment for medical error reporting and subsequent reporting of the second victim experience so that adequate support to individuals and system changes can occur. Although this study was conducted in a single institution, the structural and operational similarities between Kijabe and other tertiary hospitals in the region suggest potential applicability of findings. Further multi-site research could strengthen generalizability.

### Study Risks and Limitations

We note that recalling of adverse events can cause emotional trauma; therefore, participants were informed that they could stop participating at any time. Due to this potential, some participants quit before completing the survey. There was also a risk of participants being less than fully honest since they are employees, and discussions of the second victim experience may bring to light adverse events that could have been prevented. Additionally, the lead investigator holds a high management position. This risk was mitigated by carefully maintaining confidentiality, and the research office sent out the survey requests, and not the office of the Director of Clinical Services.

## Conclusions

Two-thirds of the healthcare workers in Kijabe reported a second victim experience. The symptoms ranged from physical symptoms, such as fatigue and sleep disturbances, to psychological symptoms, such as anxiety and depression. Counseling and psychological support were the preferred support strategies among staff members. It is important to be aware of the experiences of the second victim. Developing a support system and investing in staff well-being are important ways hospitals can support healthcare workers, which may lead to improvements in patient safety and create a more positive work environment for their healthcare providers. Hospital management have an important role in acknowledging the second victim experience and developing mechanisms to improve staff care given the high prevalence of SVE. In part, as a result of this study, the management of Kijabe Hospital initiated a staff care program that included insurance cover for external psychological counseling and support.

## Data Availability

Data is available in an online repository

https://figshare.com/s/4a7b4efd81007f000e3f

## Author contributions

**FL:** Study concept and design data analysis, data interpretation, critical revision for intellectual content, responsible for content in the final draft; **MOO**: Methodology, Writing-Original draft preparation, Investigation, Data curation, Methodology, Analysis, Writing-Reviewing and Editing

**MBA:** Study concept and design data analysis, data interpretation, critical revision for intellectual content, responsible for content in the final draft; **EN:** Data curation, Analysis, Writing-Reviewing and Editing; **PK:** Analysis, Writing-Reviewing and Editing

## Acknowledgments

We would like to express our gratitude to the dedicated staff of Kijabe Hospital for providing outstanding care for over 200,000 patients annually.

## References

1. Wu AW. Medical error: the second victim. The doctor who makes the mistake needs help too. BMJ. 2000 Mar 18;320(7237):726–7.

2. Finney RE, Torbenson VE, Riggan KA, Weaver AL, Long ME, Allyse MA, et al. Second victim experiences of nurses in obstetrics and gynaecology: A Second Victim Experience and Support Tool Survey. J Nurs Manag [Internet]. 2021 May [cited 2024 Jan 29];29(4):642–52. Available from: https://onlinelibrary.wiley.com/doi/10.1111/jonm.13198

3. Herring JA. Complications: Second Victim. J Pediatr Orthop. 2020 Jul;40 Suppl 1:S22–4.

4. Kubheka B, Naidoo S, Etieyibo E, Moyo K. Silent sufferers: Health care practitioners as second victims of patient safety incidents. Health Educ Care. 2020 Jan 1;5.

5. Sachs CJ, Wheaton N. Second Victim Syndrome. In: StatPearls [Internet]. Treasure Island (FL): StatPearls Publishing; 2024 [cited 2024 Jan 29]. Available from: http://www.ncbi.nlm.nih.gov/books/NBK572094/

6. Seys D, Wu AW, Van Gerven E, Vleugels A, Euwema M, Panella M, et al. Health care professionals as second victims after adverse events: a systematic review. Eval Health Prof. 2013 Jun;36(2):135–62.

7. Scott SD, Hirschinger LE, Cox KR, McCoig M, Brandt J, Hall LW. The natural history of recovery for the healthcare provider “second victim” after adverse patient events. Qual Saf Health Care. 2009 Oct;18(5):325–30.

8. Busch IM, Moretti F, Purgato M, Barbui C, Wu AW, Rimondini M. Psychological and Psychosomatic Symptoms of Second Victims of Adverse Events: a Systematic Review and Meta-Analysis. J Patient Saf. 2020 Jun;16(2):e61–74.

9. Vanhaecht K, Seys D, Schouten L, Bruyneel L, Coeckelberghs E, Panella M, et al. Duration of second victim symptoms in the aftermath of a patient safety incident and association with the level of patient harm: A cross-sectional study in the Netherlands. BMJ Open. 2019 Jul 1;9:e029923.

10. Chan ST, Khong PCB, Wang W. Psychological responses, coping and supporting needs of healthcare professionals as second victims. Int Nurs Rev. 2017 Jun;64(2):242–62.

11. Burlison JD, Scott SD, Browne EK, Thompson SG, Hoffman JM. The Second Victim Experience and Support Tool: Validation of an Organizational Resource for Assessing Second Victim Effects and the Quality of Support Resources. J Patient Saf. 2017 Jun;13(2):93–102.

12. Edrees HH, Morlock L, Wu AW. Do Hospitals Support Second Victims? Collective Insights From Patient Safety Leaders in Maryland. Jt Comm J Qual Patient Saf [Internet]. 2017 Sep [cited 2024 Jan 29];43(9):471–83. Available from: https://linkinghub.elsevier.com/retrieve/pii/S155372501630006X

13. Pratt S, Kenney L, Scott SD, Wu AW. How to develop a second victim support program: a toolkit for health care organizations. Jt Comm J Qual Patient Saf. 2012 May;38(5):235–40, 193.

14. Hauk L. Understanding the second victim recovery process. AORN J. 2018 Jun;107(6):P4.

15. Scott SD, Hirschinger LE, Cox KR, McCoig M, Hahn-Cover K, Epperly KM, et al. Caring for our own: deploying a systemwide second victim rapid response team. Jt Comm J Qual Patient Saf. 2010 May;36(5):233–40.

16. Wilson RM, Michel P, Olsen S, Gibberd RW, Vincent C, El-Assady R, et al. Patient safety in developing countries: retrospective estimation of scale and nature of harm to patients in hospital. BMJ. 2012 Mar 13;344:e832.

17. Nydoo P, Pillay BJ, Naicker T, Moodley J. The second victim phenomenon in health care: A literature review. Scand J Public Health [Internet]. 2020 Aug [cited 2024 Apr 21];48(6):629–37. Available from: http://journals.sagepub.com/doi/10.1177/1403494819855506

18. Strametz R, Koch P, Vogelgesang A, Burbridge A, Rösner H, Abloescher M, et al. Prevalence of second victims, risk factors and support strategies among young German physicians in internal medicine (SeViD-I survey). J Occup Med Toxicol [Internet]. 2021 Dec [cited 2024 Apr 21];16(1):11. Available from: https://occup-med.biomedcentral.com/articles/10.1186/s12995-021-00300-8

19. Rivera-Chiauzzi E, Finney Re, Riggan Ka, Weaver Al, Long Me, Torbenson Ve, Et Al. Understanding the Second Victim Experience among Multidisciplinary Providers in OBGYN. J Patient Saf [Internet]. 2022 Mar 1 [cited 2024 Apr 23];18(2):e463–9. Available from: https://www.ncbi.nlm.nih.gov/pmc/articles/PMC8521555/

20. Mathebula LC, Filmalter CJ, Jordaan J, Heyns T. Second victim experiences of healthcare providers after adverse events: A cross-sectional study. Health SA Gesondheid [Internet]. 2022 Aug 29 [cited 2024 Apr 21];27. Available from: https://hsag.co.za/index.php/hsag/article/view/1858

21. Afolalu OO, Jordan S, Kyriacos U. Medical error reporting among doctors and nurses in a Nigerian hospital: A cross-sectional survey. J Nurs Manag [Internet]. 2021 Jul [cited 2024 Jul 31];29(5):1007–15. Available from: https://onlinelibrary.wiley.com/doi/10.1111/jonm.13238

22. Mauti G, Githae M. Medical error reporting among physicians and nurses in Uganda. Afr Health Sci [Internet]. 1970 Jan 1 [cited 2024 Jul 31];19(4):3107–17. Available from: https://www.ajol.info/index.php/ahs/article/view/192294

23. Agegnehu W, Alemu A, Ololo S, Melese D. Incident reporting behaviors and associated factors among health care professionals working in public hospitals in Addis Ababa, Ethiopia 2017. MOJ Public Health [Internet]. 2019 Sep 20 [cited 2024 Jul 31];8(5):182–7. Available from: http://medcraveonline.com/MOJPH/incident-reporting-behaviors-and-associated-factors-among-health-care-professionals-working-in-public-hospitals-in-addis-ababa-ethiopia-2017.html

24. Kiguba R, Olsson S, Waitt C. Pharmacovigilance in low- and middle-income countries: A review with particular focus on Africa. Br J Clin Pharmacol [Internet]. 2023 Feb [cited 2024 Jul 31];89(2):491–509. Available from: https://bpspubs.onlinelibrary.wiley.com/doi/10.1111/bcp.15193

